# Incubation period of COVID-19 in the “live-house” cluster of accurately known infection events and delay time from symptom onset of public reporting observed in cases in Osaka, Japan

**DOI:** 10.1101/2020.05.23.20110908

**Authors:** Toshihisa Tomie

## Abstract

The incubation period of an infectious disease is very important for control of the disease but estimating the period is not easy because the date of infection is not easy to identify. Accurate incubation period distribution by examining cases in the cluster generated in “live-houses” in Osaka, Japan with known infection events is reported. The distribution of the latent period is also estimated. The modes of incubation and latent periods of COVID-19 in Japan are 4.1 days and 3.3 days, respectively. The mode of the delay time from the onset to reporting is estimated to be 4.7 days, telling that the effects of interventions show up in the number of infections two weeks later after the measures.

## 1. Introduction

The incubation period of an infectious disease is the time between infection and symptom onset. This period is very important for surveillance and control of the disease because symptom onset is the only indication of the infection before nation-wide investigations, such as, by PCR (polymerase chain reaction) test of the virus when the infection is caused by a virus. By knowing the distribution of the period, public-health officials can set the quarantine period of infected individuals to suppress the epidemic of the disease in the society without the aid of a vaccine for the virus.

However, estimating the incubation period is not easy because the date of infection is not easy to identify. There are many reports^1-7)^ on the incubation period of the COVID-19 virus after its outbreak in Wuhan, China in January 2020. In ref. 4, 106 cases of COVID-19 were analyzed and the number of cases is large enough to discuss statistics. But the accuracy of the date of contact with the infection source was three days, which is not sufficient for assessing the incubation period of several days. In refs. 1, 2, 3, and 5, the date of infection was estimated from the period of stay in Wuhan, which will be also not accurate enough. In ref 6, the incubation period of COVID-19 was evaluated from the onset date of 5 persons who had dinner with a primary infected person. Reference 7 estimated the incubation period from the onset date of four Germans who had a business meeting with a visitor from China. In both references, although the date of infection was clear, the number of cases studied was too small to discuss statistics.

In this paper, we report accurate incubation period distribution by examining 48 cases in a cluster generated in “live-houses” in Osaka Prefecture in Japan with accurately known infection events. The distribution of the latent period of infection, which is the time from infection to onset of infectiousness, was also obtained from the difference in the onset dates of the secondary and the primary infection from 64 pairs reported by Osaka Prefecture.

We also report the distribution of the delayed days from onset to reporting estimated from more than 1,700 cases reported by Osaka Prefecture. An infected person does not go to a medical institution immediately after his onset. It takes time to get PCR test results in medical institutions. It also takes time to collect data by a local government to report public. Very often, people use the daily reported number of infections rather than the daily change of the number of people on the date of infection. Therefore, we need to know also the distribution of delayed days from the onset of the disease to the reporting by a local government for assessing the effect of any human interventions to control infectious diseases.

## 2. Method

Several COVID-19 infection clusters have been observed in Japan. Especially in Osaka Prefecture, 48 infected people of “live-house” spectators have been confirmed^8)^. “Live-house” is a small venue where a small concert is organized with spectators of about 100 people with small meals serviced often without seats. The dates of the live-house events to which infected people attended were precisely recorded. Records of people close contacted with the primary infects who were infected at “live-houses”, have also been made public. From the delay of the onset date of the secondary infection to the onset date of the primary infection, we can evaluate the latent period from the infection to the day obtaining infection power by assuming the same incubation period from infection to onset for both of primary and secondary infections.

Osaka Prefecture has announced more than 1,700 cases of infection, and the date of onset is known for most cases. From this data, the number of days delayed from onset to reporting can also be assessed. Some of cases are documented to have close contact with infected people.

In a historical paper^9)^, Sartwell first showed that the incubation period distribution of many infectious diseases from a point source followed “log-normal” distributions, which were confirmed in refs.10 and 11 and in many other papers cited in ref.12. We apply the Sartwell model and fitted the observed incubation and latent period distributions by log-normal distributions. The distribution of delayed days from onset to reporting was also well described by a log-normal distribution.

## 3. Results

### 3-1. Incubation period; the time from infection to onset

A COVID-19 infection cluster occurred in four “live-houses” in Osaka. The first infectious case was reported on February 27 and the last case was found on March 12, lasting two weeks.

Concerts were held on two days at venue A on February 15 and 16, on three days at B venue on 19, 23, and 24, one day at C venue on 18, and one day at D venue on 21. Of those who participated as spectators or worked as staff, 48 people were identified to be positive for COVID-19 virus by the PCR test.

Although some people participated in the concerts more than once, many of them participated only once, so the dates of infection are accurately identified. Some positive for PCR did not develop symptoms. Of those who attended the concert only once and were able to identify the date of infection event, a total of 24 cases were infected and showed symptoms.

The distribution of the incubation period (the number of days from the day of participating in the concert to the day of onset) of these persons is shown in Fig. 1. Day 0 is the day when an infected person attended a concert.

**Fig. 1:**
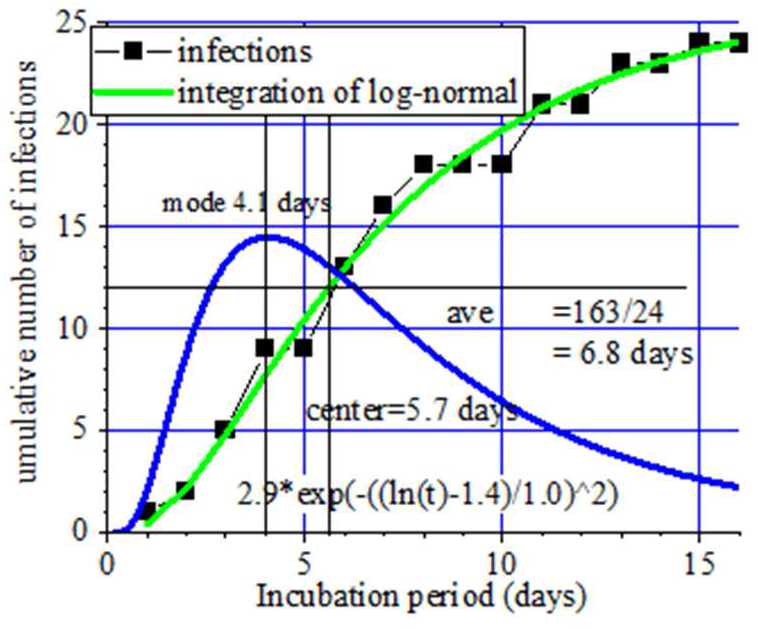
Distribution of incubation period.

Since the statistical noise of the number of cases in a single day is large, the fitting was performed to the history of the cumulative number of cases with a log-normal equation, and the result was 2.9 × exp (-((ln (t)-1.4) /1.0)^2^).

The most frequent incubation period, mode, was 4.1 days, the median was 5.7 days, 10% -90% values were 2 days-12 days, and the average period was 6.8 days.

### 3-2. Latent period; the time from infection to onset of infectiousness

Of 346 cases reported by April 3 in Osaka Prefecture, 89 cases were reported to have close contact with an infected person. Among 89 cases, 64 pairs had records of onset dates for both infection and infector. Figure 2 shows the distribution of frequency of pairs as a function of days of the clinical onset of the secondary infection from that of the primary infection. The distribution is well described by a log-normal, 11 × exp (-(ln (t)-1.2) /0.8)^2^).

**Fig.2;.**
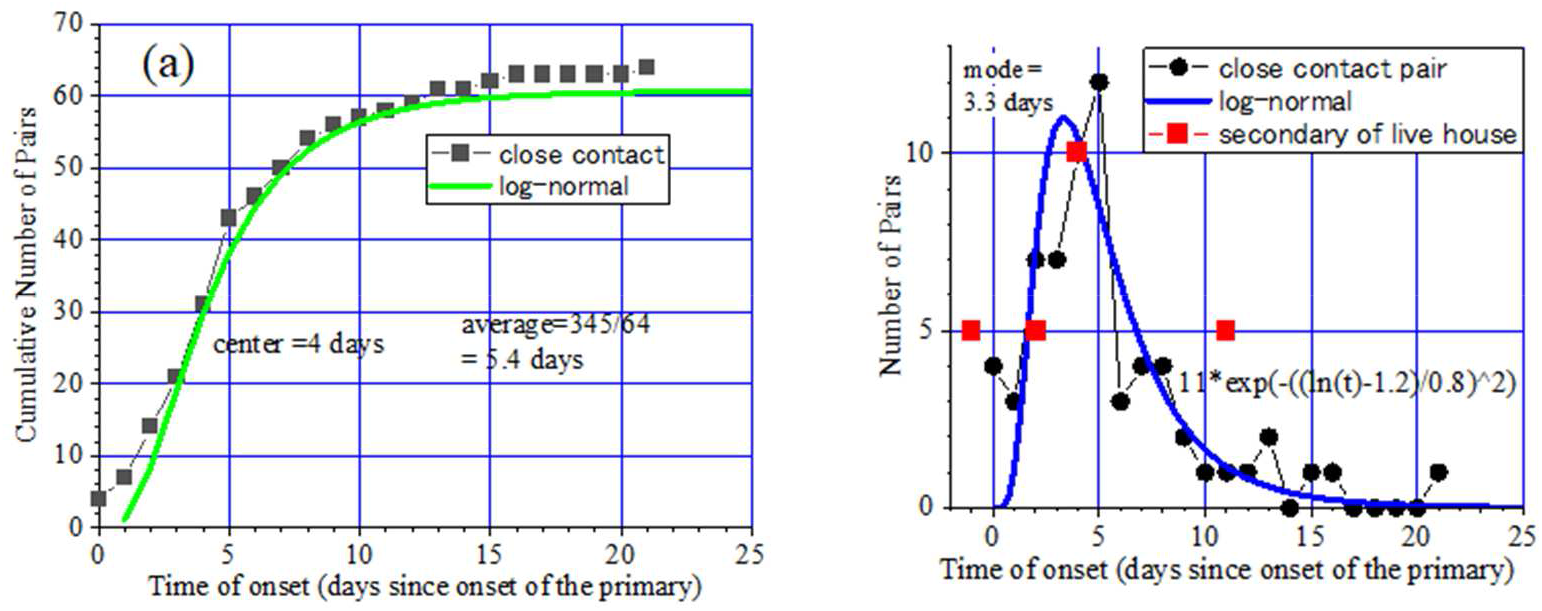
Cumulative number (a) and number in a single day (b) of pairs as a function of time of onset of the secondary infection after that of the primary.

The mode was 3.3 days, the median was 4 days, 10% -90% was 2.5days -8 days, and the average latent period was 5.4 days.

Of the close contacts of those infected in the live-house, 23 persons were positive. Among them, 5 pairs had a record of onset. The distribution of these 5 pairs are shown by red squares followed fairly well the above log-normal curve.

The difference in onset time between infections and infectors is usually called “serial interval”^13)^. But in the cases of the coronavirus epidemic in Osaka, except in the cluster of “live-house”, there is no clear history of being infected for all of the primary infections. We just assumed that a person who showed symptom earlier in a pair was the primary, but we are not sure whether the “assumed” secondary infection was really infected from the “assumed” primary infection. Moreover, there were very few cases of having a tertiary infection. So, we are reluctant to use the terminology of the “serial” interval in our analysis.

When the clinical onset is *T_onset_* and the time of transmission to other people from infection, latent period, is *T_trans_*, the difference of dates of onset of person A and B when B is the secondary, *T_AB_*, is given by *T_AB_= T_onset,A_+ T_transs_* - *T_onset,B_*. If *T_onset,A_= T_onset,B_*, then, *T_AB_= T_trans_* which is the latent period.

### 3-3. Delayed days from onset to reporting

In Osaka, 1,743 cases were reported by May 10. Although some people are asymptomatic or have unknown onset dates, most people's onset dates have been announced. Figure 3 shows the distribution of the number of delayed days from the onset date to the reporting date.

**Fig.3:**
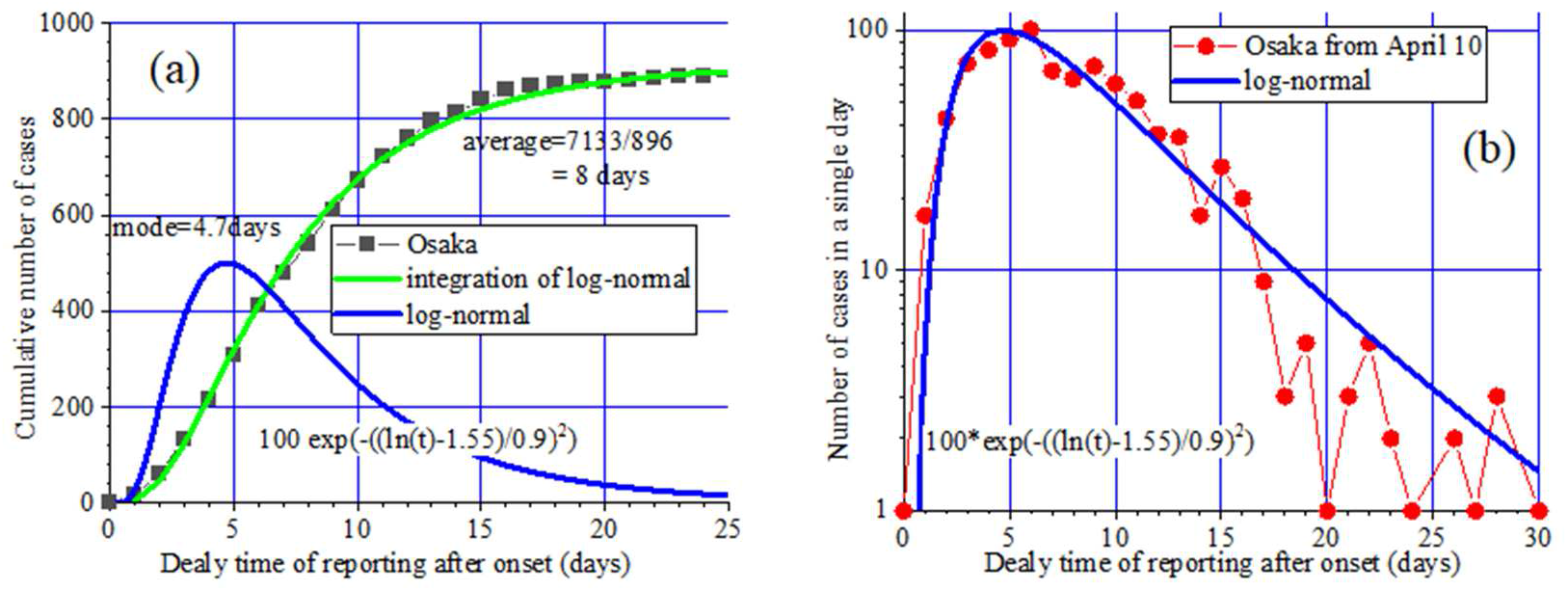
Cumulative number (a) and number in a single day (b) of reported cases as a function of delayed time of reporting after onset.

Fitting the history of the cumulative number of cases with a log-normal distribution gives, 100 × exp (-((ln (t) -1.55) /0.9)^2^).

The mode was 4.7 days, the median was 7 days, and the average was 8 days.

## 4. Discussions

Since the average value of the incubation period is 6.8 days and the average number of days delayed from the onset to the announcement is 8 days, the number of infected persons reported is information two weeks before. If human interventions to a disease can change the infection immediately, for example, if the infection rate can be immediately halved, the number of infected people will be halved two weeks after the measures unless other conditions change.

The mode of the incubation period of 4.1 days is shorter than the mode of the latent period of 3.3 days, which means that the infectivity is maximum at 1 day before the onset of symptoms. Interestingly, Fig. 2 (a) shows that 9 days after infection, 90% of infections has lost the infectivity, whereas Fig. 1 shows that at this time, 30% of people did not show symptom. We could say that, at the time of onset, the infectivity is already lost. If this is the case, it would be meaningless to isolate the infected person for suppressing an epidemic, which is unconvincing.

As the incubation period of SARS Coronavirus, ref. 8 reported 4 days, refs. 3 and 5-7 reported 5 days, ref. 4 reported 6.4 days, and ref.1 reported 10 days. This variability of values in literature might be primarily due to inaccurate estimates of infection dates.

There are many reports of pre-symptomatic transmission of VID-19. Ref. 14 reported that the primary and the secondary infections showed symptoms on the same day in two clusters among 5 clusters they studied. Reference 4 reported 4 secondary cases among 74 secondary cases showed symptoms before that of the first generation. The first generation had a history of visiting Wuhan while the secondary did not.

In our case, in the cluster of “live-house”, a wife who was infected by her husband who was a spectator in a “live-house” showed symptom one day earlier than her husband as a primary infection, as is shown by the red square at the time of -1.

The fact that the mode of the latent period shown in Fig.2 is smaller than the mode of the incubation period shown in Fig. 1 means that most of the transmission occurred before the onset of the primary. It is not easy to understand what can cause the dispersion of the latent period distribution narrower than that of the incubation period.

## 5. Summary

By examining COVID-19 clusters created in “live-houses” in Osaka Prefecture in Japan with well-documented infectious events, we obtained an accurate distribution of the latent period. The distributions of the incubation and latent periods, and the number of delayed days from the onset to the report all followed log-normal distributions and they are exp (-((ln (t) -1.4) /1.0)^2^), exp (-((ln (t) -1.55) /0.9)^2^), and exp (-((ln (t) -1.55) /0.9) ^2^), respectively. The shorter latent period than the incubation period implies that transmission of the pathogen takes place before the symptom. The sum of the averages of the incubation period and the delayed days from the onset to reporting is about two weeks, telling that the effects of interventions show up in the reported number of cases two weeks after the measures.

## Data Availability

All data referred are available from references in the manuscript.

## References

1. N. M. Linton, T. Kobayashi, Y. Yang, K. Hayashi, A.R. Akhmetzhanov, S. Jung, B. Yuan, R. Kinoshita, and H. Nishiura, “Incubation Period and Other Epidemiological Characteristics of 2019 Novel Coronavirus Infections with Right Truncation: A Statistical Analysis of Publicly Available Case Data.” J Clin Med. 9, (2020); [PMID: 32079150] doi:10.3390/jcm9020538

2. J.A. Backer, D. Klinkenberg, J. Wallinga “Incubation period of 2019 novel coronavirus (2019-nCoV) infections among travellers from Wuhan, China, 20-28 January 2020”. Euro Surveill. 25 (2020). [PMID:32046819] doi:10.2807/1560-7917.ES.2020.25.5.2000062

3. Q. Li, X. Guan, P. Wu P, et al. “Early transmission dynamics in Wuhan, China, of novel coronavirus-infected pneumonia.” N Engl J Med. 2020. [PMID: 31995857] doi:10.1056/NEJMoa2001316

4. W. Xia, J. Liao, C. Li, Y. Li, Xi Qian, X. Sun, H. Xu, G. Mahai, X. Zhao, L. Shi, J. Liu, Yu, M. Wang, Q. Wang, A. Namat, Y. Li, Ji. Qu, Qi Liu, X. Lin, S. Cao, S. Huan, J. Xiao, F. Ruan, H. Wang, Q. Xu, X. Ding, X. Fang, F. Qiu, J. Ma, Y. Zhang, A. Wang, Y. Xing, S. Xu, “Transmission of corona virus disease 2019 during the incubation period may lead to a quarantine loophole” Preprint medRxiv doi:10.1101/2020.03.06.20031955

5. S. A. Lauer, K.H. Grantz, Q. Bi, Forres t K. Jones, Qulu Zheng, H. R. Meredith, Andrew S. Azman, N. G. Reich, and J. Lessler, “The Incubation Period of Coronavirus Disease 2019 (COVID-19) From Publicly Reported Confirmed Cases: Estimation and Application” Ann. Intern. Med. 122, (2020) 577. doi:10.7326/M20-0504

6. Y. Gao, C. Shi, Y. Chen, P. Shi, J.Liu, Y. Xiao, Y. Shen, E. Chen, “A cluster of the Corona Virus Disease 2019 caused by incubation period transmission in Wuxi, China” J. Infection 80, (2020), 666.

7. C. Rothe, M. Schunk, P. Sothmann, G.Bretzel, G. Froeschl, C. Wallrauch, T. Zimmer, V. Thiel, C. Janke, “Transmission of 2019-nCoV Infection from an Asymptomatic Contact in Germany”. N Engl J Med. Published online Jan. 30, 2020. doi:10.1056/NEJMc2001468

8. http://www.pref.osaka.lg.jp/hodo/index.php?HST_BUC0DE=6000&HST_SY0C0DE=&HST_STARTDATE2_START=&HST_STARTDATE2_END=&HST_STARTDATE2=&HST_TITLE1=%90V%8C%5E%83R%83%8D%83i%83E%83C%83%8B%83X&SEARCH_NUM=50&searchFlg=%8C%9F%81%40%8D%F5&site=fumin&start=3

9. P.E. Sartwell, “The Distribution of Incubation Periods of Infectious Diseases” Am. J. Hygiene (1950) 51 pp.310–18 (reprinted in Am. J. Epidemiol. 141, (1995) 386.

10. J. Lessler, T. M. Perl, K. E. Nelson, D. A. T. Cummings, N. G. Reich, R. Brookmeyer, T. M. Perl, “Incubation periods of acute respiratory viral infections: a systematic review” Lancet Infect Dis. 9, (2009) 291. doi:10.1016/S1473-3099(09)70069-6

11. R. González-Riveral, R.C. Culverhouse, A. Hamvas, P.I. Tarr, and B.B. Warner, “The age of necrotizing enterocolitis onset: an application of Sartwell’s incubation period model” J. Perinatology 31, (2011) 519.

12. P. E. M. Fine, “The Interval between Successive Cases of an Infectious Disease” Am. J. Epidemiol. 158, (2003) 386.

13. E. Kenah, M. Lipsitch, and J. M. Robins, “Generation interval contraction and epidemic data analysis” Math. Biosci. 213, (2008), 71.

14. W. E. Wei, Z. Li, C. J. Chiew, S. E. Yong, M. P. Toh, V. J. Lee, “Presymptomatic Transmission of SARS-CoV-2 — Singapore, January 23–March 16, 2020”, Morbidity Mortality Weekly Report, 69, (2020)411.

